# Facility-level variation in HIV service delivery volumes before and after the 2025 PEPFAR funding cuts: a retrospective analysis of District Health Information System data in Zambia

**DOI:** 10.64898/2026.09.27.26364124

**Authors:** Mike Mwale, Sydney Rosen, Thandiwe Ngoma, Suilanji Sivile, Chimuka Sianyinda, Prudence Haimbe, Hilda Shakwelele, Mariet Benade, Allison Morgan, Amy Huber, Idah Mokhele

## Abstract

**Background:** The abrupt withdrawal and uneven restoration of United States PEPFAR support in 2025 disrupted HIV service delivery in aid-dependent countries worldwide. In Zambia, decreases in HIV service delivery have been documented at a national level, but variations by facility and by quarter have largely been masked by existing aggregate district– and province-level estimates. We examined facility-level variation in HIV service volumes delivered before and after the funding cuts.

**Methods:** We analysed monthly District Health Information System (DHIS2) data covering testing volumes, ART initiations, and numbers on ART for January 2024-December 2025) for two sets of facilities: 1) 24 selected SENTINEL network facilities across four provinces (Central, Copperbelt, Lusaka, and Southern); and 2) all facilities in two districts, Chongwe in Lusaka Province, which was supported by the U.S. CDC under PEPFAR, and Mpongwe in Copperbelt Province, which was supported by USAID under PEPFAR. For each facility, we calculated quarterly 2025:2024 ratios of service volumes and summarized quarterly and annual percentage changes in numbers of tests, treatment initiations, and clients on ART.

**Results:** Aggregate HIV testing volumes declined 18% in SENTINEL facilities from 2024 to 2025, with 17 of 24 facilities showing net annual decreases. Aggregate ART initiations declined 16%, with 23 of 24 facilities experiencing net decreases. For both indicators, variation between quarters and among individual facilities matched or exceeded aggregate changes. Numbers of clients currently on ART were comparatively stable, with only 6 facilities showing net declines exceeding 5%. District-level analyses showed divergent patterns. Net aggregate testing volumes increased by 7% in Chongwe District, but 28/39 facilities experienced quarterly declines exceeding 10% in at least one quarter and fell by 40% in Mpongwe District overall. Only 4 of 36 sites in Chongwe showed a net loss of clients over the course of 2025, with most experiencing gains, while Mpongwe experienced substantial variation by facility, with many having declines of >30% during some quarters and 14 of the 25 facilities suffering a net loss from 2024 to 2025.

**Conclusions:** Facility-level performance varied substantially within the same district following the 2025 PEPFAR funding disruptions, with some sites losing most of their service volumes while others remained stable or exceeded 2024 levels. These findings emphasize the need for facility-level monitoring to target recovery support where it is needed most.

## Introduction

In 2024, the U.S. President’s Emergency Plan for AIDS Relief (PEPFAR) funded 84% of the national HIV program budget of Zambia, a high HIV prevalence country in southern Africa[1]. PEPFAR funding was channeled via two main United States Government (USG) agencies, USAID and the CDC. Funds were then distributed to governmental and nongovernmental “implementing partners” (IPs), which delivered both technical assistance, such as coordination, laboratory capacity, and policy development, and direct services to healthcare facilities and in communities, such as personnel, equipment, and commodities, in close collaboration with Zambia’s Ministry of Health. Zambia reported exceeding all three UNAIDS “95-95-95” targets for testing, treatment, and viral suppression by the end of that year [2].

On January 24, 2025, the USG issued a “stop work order” that brought all PEPFAR activities to an immediate halt worldwide. IP assistance to healthcare facilities was abruptly frozen for several weeks in February and March 2025, following the stop work order. Although support gradually returned at some facilities for “lifesaving” services (primarily HIV testing and treatment[2]), most USAID-funded IP contracts were terminated at least temporarily, as was funding for ancillary CDC-supported services, such as data systems and laboratory sample transport [1,3].

Over the course of 2025, assistance to healthcare facilities from all IPs fluctuated frequently, with some former USAID IPs receiving funding from the U.S. Department of State to sustain some programs and many IPs receiving short-term contract extensions that were renewed unpredictably and with or without additional funding[4]. At the facility and community levels, IP staff and other resources were withdrawn and restored intermittently, in accordance with funding availability and USG instructions. In response, the Zambian Ministry of Health (MOH) redeployed government staff to healthcare facilities to maintain delivery of core HIV services, such as antiretroviral therapy (ART) provision, but it was not able fully to replace the estimated 11,000 healthcare staff previously supported by PEPFAR [5].

Since the 2025 stop work order, the impact of the loss of PEPFAR support on Zambia’s HIV program has been described or modeled several times[1,6–9]. Most of these efforts have addressed high-level losses, based either on data from the national HIV electronic medical record system or on mathematical models, and report average or aggregate results. While critical for understanding overall changes, these aggregate analyses mask variations in on-the-ground impacts of the service delivery cuts at the level of individual healthcare facilities. If individual healthcare facilities are affected by external funding cuts differently, even within the same subnational geographic districts, then tailoring resource allocation and interventions to match the degree of impact may be a more effective response strategy than blanket national policy responses.

To understand facility-level experiences, we observed service delivery at a network of selected facilities that we have monitored for several years under other studies[10,11]. We report facility-level data on HIV test volumes, treatment initiations, and numbers currently on ART from a sample of healthcare facilities in four of Zambia’s 10 provinces and concentrated analysis of HIV testing numbers for all facilities within two districts between January 2024 and December 2025.

## Methods

### Study sites and data

This analysis uses monthly data from the District Health Information System 2 (DHIS2) from January 2024 to December 2025. DHIS2 is Zambia’s national routine health information system, which aggregates facility-level service delivery data on a monthly basis[12]. HIV data for DHIS2 are generated from paper-based facility registers and from the national electronic medical record system, known as SmartCare Pro, which serves more than 90% of clients on ART[13]. We accessed DHIS2 data from the Ministry of Health’s Health Management Information System (HMIS) national portal.

Data were extracted for two sets of study sites, as illustrated in Figure 1. The first (Dataset 1) comprised the 24 facilities across four provinces–Lusaka, Southern, Central, and Copperbelt– included in the SENTINEL network, which has been under observation by the study team since 2018 (Table 1)[10]. Six sites from each province were chosen for study participation and aimed to capture variation in setting (urban vs rural), PEPFAR support (USAID vs CDC), and differentiated service delivery model offerings. They now comprise a network of facilities at which new issues of public health importance, such as the impact of global funding cuts, can be assessed. All are relatively large by ART patient volume; some are attached to hospitals, and some are co-managed by the Ministry of Health and local religious missions. Table 1 also indicates PEPFAR support that was (or is) provided by an implementing partner. As noted above, the two funding agencies utilized different funding models: CDC supported provincial health offices, which were then responsible for implementing the agreed-upon HIV services, while USAID directly funded nongovernmental implementing partners.

**Figure 1.**
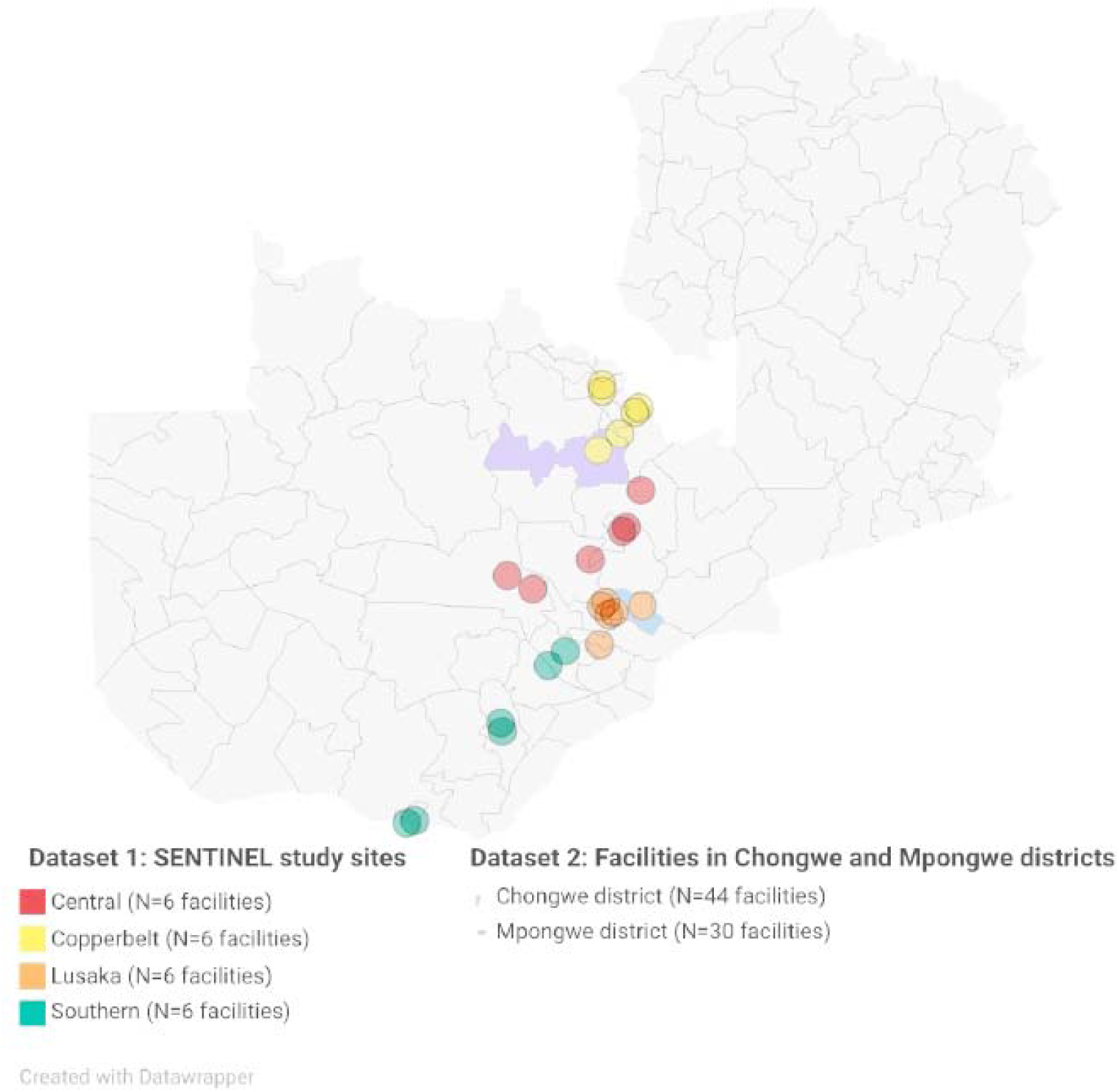
Locations of study sites.

**Table 1.** SENTINEL study sites (Dataset 1)

| Province | Site | Setting | Level | Funder and past or current role of IPs |
| --- | --- | --- | --- | --- |
| Lusaka | L1 | Urban | Urban health centre | CDC; IP for technical assistance and Ministry of Health (Lusaka Provincial Health Office) for implementation support |
|  | L2 | Rural | Rural health centre |  |
|  | L3 | Urban | Health centre |  |
|  | L4 | Urban | First level hospital |  |
|  | L5 | Urban | First level hospital |  |
|  | L6 | Urban | Urban health centre |  |
| Southern | S1 | Urban | Urban health centre | CDC; Ministry of Health (Southern Provincial Health Office) for technical assistance and implementation support |
|  | S2 | Urban | Urban health centre |  |
|  | S3 | Urban | Urban health centre |  |
|  | S4 | Urban | Urban health centre |  |
|  | S5 | Urban | General hospital |  |
|  | S6 | Urban | Urban health centre |  |
| Central | C1 | Rural | District hospital | USAID; IP for technical assistance and implementation support |
|  | C2 | Urban | Urban health centre |  |
|  | C3 | Urban | Urban health centre |  |
|  | C4 | Urban | Urban health centre |  |
|  | C5 | Rural | Mission hospital |  |
|  | C6 | Rural | District hospital |  |
| Copperbelt | B1 | Urban | Urban health centre | USAID; IP for technical assistance and implementation support |
|  | B2 | Urban | Urban health centre |  |
|  | B3 | Rural | Mission hospital |  |
|  | B4 | Rural | Mission hospital |  |
|  | B5 | Urban | Urban health centre |  |
|  | B6 | Urban | Urban health centre |  |

The second set of study sites (Dataset 2) comprised all public and private sector healthcare facilities recognized by the Ministry of Health in Chongwe District in Lusaka Province and Mpongwe District in Copperbelt Province. Chongwe district contains one SENTINEL site and Mpongwe district contains two SENTINEL sites. The districts were funded under different PEPFAR funding mechanisms. In addition to the same DHIS2 dataset cited above, we also utilized the Ministry of Health’s Master Facility List to identify the level of each facility[14].

The facilities included in Dataset 2 for each district are briefly described in Table 2. Throughout this paper, each site is identified by a code consisting of a letter for the province or district and a number for the site (e.g. “L1” for the first site in Lusaka Province); the codes are arbitrary but allow results for the same facility to be compared for the three indicators.

**Table 2.** Facilities in Chongwe and Mpongwe districts included in the analysis (Dataset 2)

| Variable | Chongwe District | Mpongwe District |
| --- | --- | --- |
| PEPFAR partner agency | CDC | USAID |
| Total number of facilities | 43 | 30 |
| Setting |  |  |
| Urban | 4 | 0 |
| Rural | 39 | 30 |
| Level |  |  |
| Health post | 18 | 14 |
| Health Centre | 22 | 14 |
| First level hospital | 2 | 2 |
| Third level hospital | 1 | 0 |

### Data analysis

To understand changes in HIV testing and treatment initiations, we compared quarterly totals of the volumes recorded for each study site before and after the funding cuts, reporting the ratio of 2025 (after) to 2024 (before) values (e.g. total ART initiations at a study site in quarter 1 of 2025 divided by total ART initiations at the same facility in quarter 1 of 2024). To understand fluctuations in the number of clients on ART, we calculated the quarterly average number of individuals on ART at each study site and compared the average values before and after the funding cuts. Ratios above 100% indicate that service volumes in 2025 exceeded those recorded in the corresponding quarter of 2024, while ratios below 100% indicate lower service volumes relative to the same period in 2024. For numbers of clients on ART, we used quarterly averages of persons in care and report the same ratios.

For presentation purposes, ratios were colour-coded into five bands to show the scale of divergence from 2024 levels: dark red (<50%: 2025 values were substantially lower than 2024 values), light red (50– 75%, moderately below), amber (76–100%, 2025 values were broadly comparable to 2024), light green (101–129%, moderately above), and dark green (≥130%, substantially above).

We also calculated total and percentage changes in annual testing and ART initiation volumes and in the annual average number of persons in care at each facility over each 12-month period to estimate net differences between 2024 and 2025. For the district facility set analysis, we analyzed only HIV testing volumes and current numbers on ART, as ART initiation data were incomplete.

Facilities in both datasets had missing DHIS2 data in several domains, though missing entries for SENTINEL sites (Dataset 1) were much less frequent than for the district facility set (Dataset 2). For HIV testing volumes, where either year (2024 or 2025) was missing data for a given month, both years’ values for that month were excluded (paired exclusions), so that quarterly totals always compared the same set of months in each year. For current numbers on ART, quarterly results were averages of the monthly values available for that quarter. For example, if numbers on ART were only available for two of the three months in a quarter, we averaged the two available data points and multiplied by three to represent the full quarter. For all indicators, sites missing data for more than three different calendar months during the two-year study period were excluded from the analysis entirely, to avoid having to exclude more than 25% of the months in either 12-month year. Months in which data were missing and the number of months contributing data for each site and quarter are shown in Supplementary Table 1.

### Artificial intelligence (AI) disclosure statement

ChatGPT 5.6 Sol was utilized during the preparation of this manuscript to design heatmap figures from data provided, check internal consistency of data and results reported, edit text for clarity, copyedit text, identify some citations and design the tables. The authors collected and analyzed all data, reviewed all AI contributions, and confirmed all citations. The authors take full responsibility for the accuracy, integrity, and final content of this publication.

Ethics

All data used in this analysis were aggregate, non-human subjects’ data. The study team had no interaction with any individual research participants. Access to DHIS2 data was approved by the ERES Converge IRB through the GREAT protocol (2019-Sep-030) and the SHIFT protocol (2025-Sep-011). Both the GREAT and SHIFT protocols were also approved by the National Health Research Authority (NHRA) in Zambia and by the Boston University Medical Campus IRB (GREAT: H-38823; SHIFT: H-46287).

## Results

### HIV testing at the SENTINEL sites

Figure 2 presents the ratio of the quarterly number of HIV tests at each SENTINEL site in 2025 to tests conducted in 2024. It also describes the absolute and percentage change in total annual tests completed in 2024 and 2025. For example, at site L1, the volume of HIV tests performed in the first quarter of 2025 was 28% higher than the volume of tests performed in the first quarter of 2024. This site continued to complete more tests in the next two quarters of 2025 but then saw a 7% absolute reduction below 2024 levels in quarter four. In aggregate, a total of 15% more tests were performed at L1 in 2025 than in 2024. L3, in contrast, experienced a 22% net decline in the number of tests performed, despite increases in the first two quarters of 2025.

**Figure 2.**
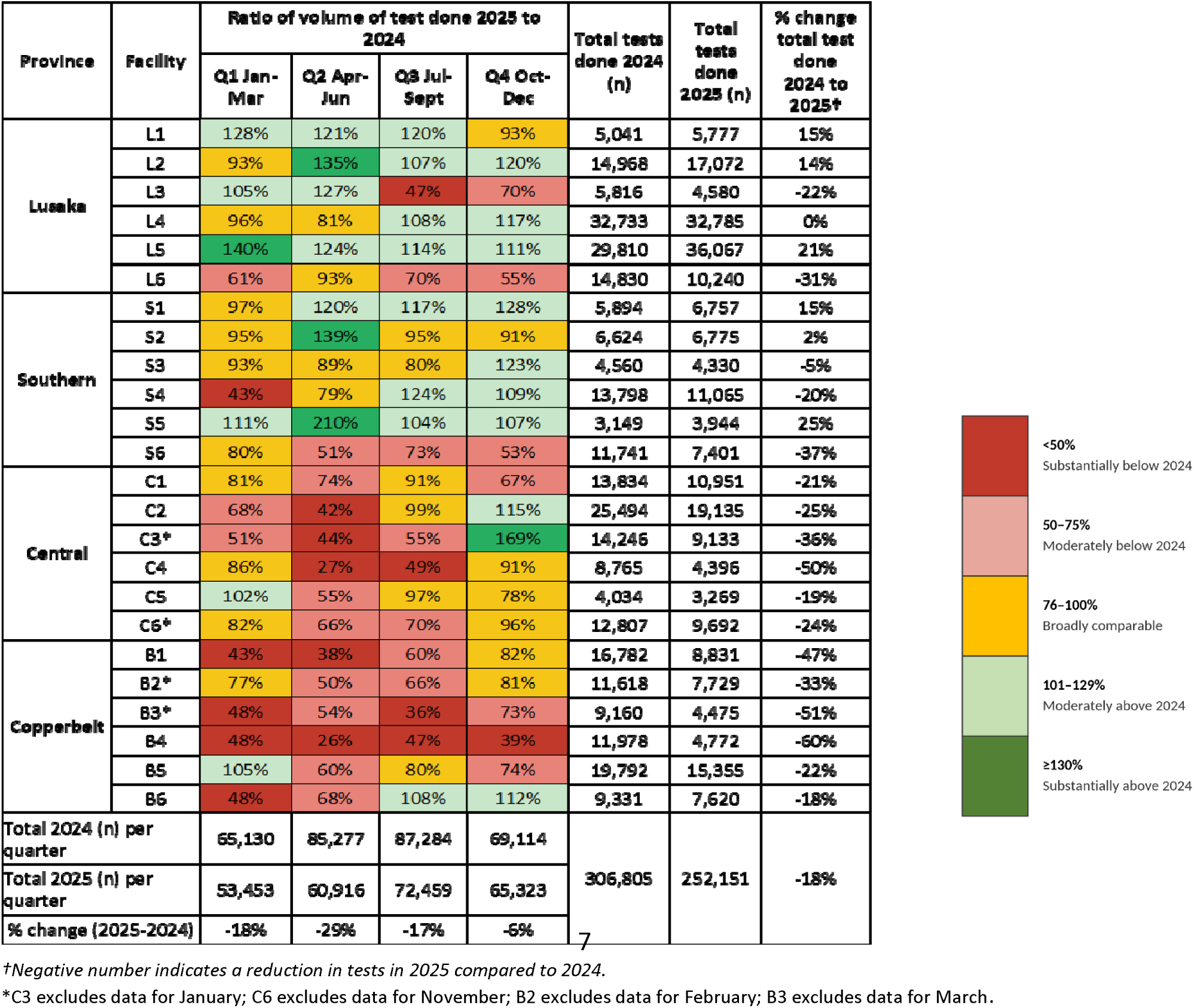
Quarterly ratios and total annual numbers of HIV tests conducted in 2025 relative to the corresponding interval in 2024 at 24 SENTINEL sites, by facility.

As illustrated in Figure 2, numbers of HIV tests conducted at each study site fluctuated widely between quarters. Facilities in Lusaka Province generally maintained testing volumes at or above 2024 levels, though testing volumes at sites L3 and L6 fell off sharply during the second half of 2025. Like Lusaka Province, facilities in Southern Province generally maintained testing volumes between the years, though with some experiencing declines in some quarters.

Testing volumes broadly decreased across Central Province, where most facilities showed steep declines in quarter two in particular. Recovery at the Central Province sites by the end of 2025 was uneven. Facilities in Copperbelt Province experienced the largest and longest decline, with the majority reporting testing volumes below 70% of 2024 levels for several consecutive quarters.

The percentage change in total annual tests conducted in 2024 and 2025 ranged from an increase of 25% (S5 in Southern Province) to a decrease of 60% (B4 in Copperbelt Province); 17 of the 24 facilities experienced a net decrease in testing volumes from 2024 to 2025, leading to a decrease of 18% for the SENTINEL sites as a whole. Interestingly, Figure 2 also suggests that variability among sites within districts is comparable in magnitude to variability between districts, indicating that aggregate estimates may mask important local differences in performance.

### ART initiations at the SENTINEL sites

Figure 3 illustrates quarterly differences in numbers of clients initiated on ART at each facility between 2024 and 2025. Most facilities in all provinces experienced meaningful declines in at least two quarters of 2025, relative to 2024. Patterns of increases and decreases in ART initiations vary less strongly by province than did HIV testing volumes, though facilities in Lusaka and Southern provinces again had fewer quarterly declines than did Central and Copperbelt provinces. With the notable exception of L2, all study sites conducted fewer ART initiations in 2025 than in 2024, resulting in an overall reduction of 16% for the full network of SENTINEL sites.

**Figure 3.**
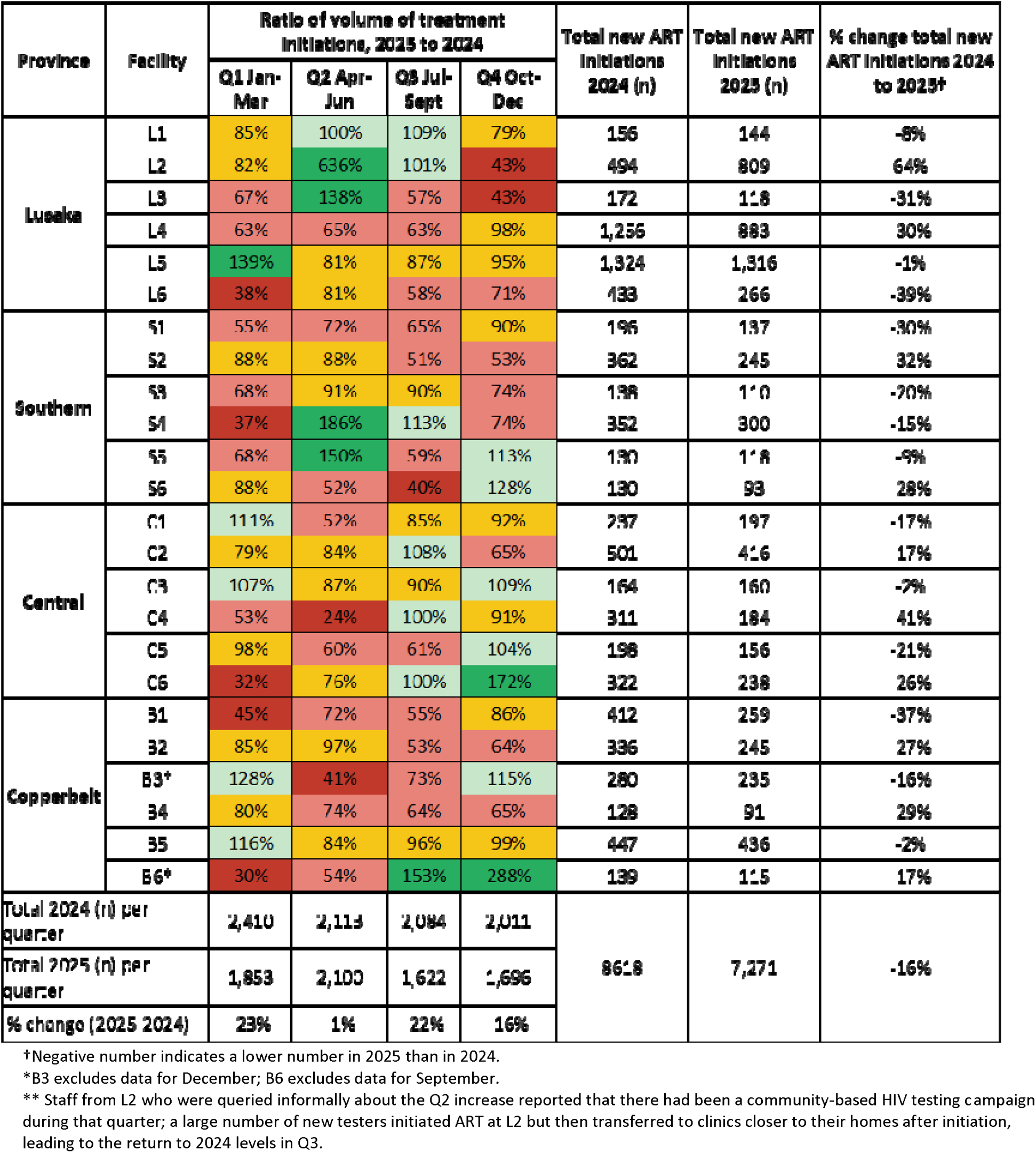
Quarterly ratios and total annual numbers of ART initiations in 2025 relative to the corresponding interval in 2024 at SENTINEL sites, by facility.

### Numbers of clients on ART at the SENTINEL sites

In Figure 4 we present DHIS2 data on the average number of ART clients at each SENTINEL site, by quarter and year. ART numbers were much more stable over the study period than were numbers of HIV tests or ART initiations, with most facilities remaining within 20% of baseline throughout the period. Facilities in Copperbelt Province were most likely to dip below this ratio. Annual averages also remained quite stable, with only 6 facilities ending 2025 with a net reduction in ART numbers of greater than 5%. Three of these facilities were in Copperbelt Province.

**Figure 4.**
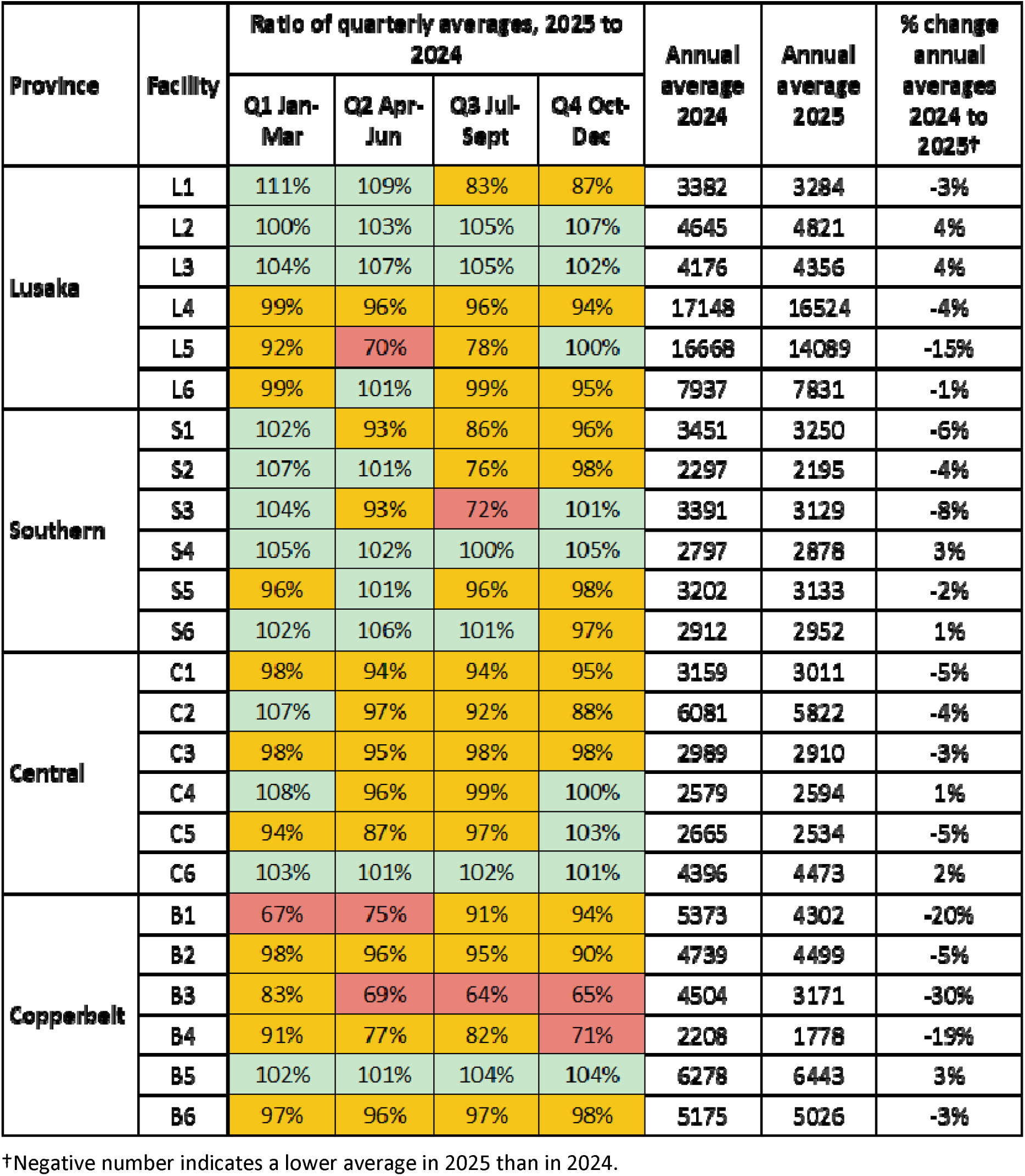
Ratios of quarterly averages and annual average numbers on ART in 2025 relative to the corresponding interval in 2024 at SENTINEL sites, by facility.

### HIV testing in Chongwe and Mpongwe districts

To discern the extent to which individual facility losses or gains reflect movement of clients between facilities within districts, we analyzed HIV testing volumes in all facilities included in DHIS2 in Chongwe (n=44) and Mpongwe (n=30) districts. After removing facilities that were missing too many quarterly observations in the database to allow analysis, the analytic dataset for HIV testing included 39 facilities in Chongwe and 27 in Mpongwe.

In Chongwe District (Figure 5), while testing volumes fluctuated widely, both between quarters and among sites, by quarter 4 of 2025, 41% of the facilities were conducting as many as or more HIV tests than they had in the previous year, though numbers at a few (e.g., CF16, CF17, CF18) had dropped off sharply. One relatively small facility, CF4, had unusually high ratios in Q1 and Q3 compared to the other facilities, for reasons that we could not learn. The overall percentage change in total tests conducted across the 39 facilities ranged from a 36% decline to a 237% increase between 2024 and 2025, with just over half of facilities reporting a net increase in annual testing volume. Despite some quarters in which some facilities conducted fewer than half the anticipated number of tests, the net difference in the number of HIV tests performed in Chongwe District between 2024 and 2025 was a positive 7%.

**Figure 5.**
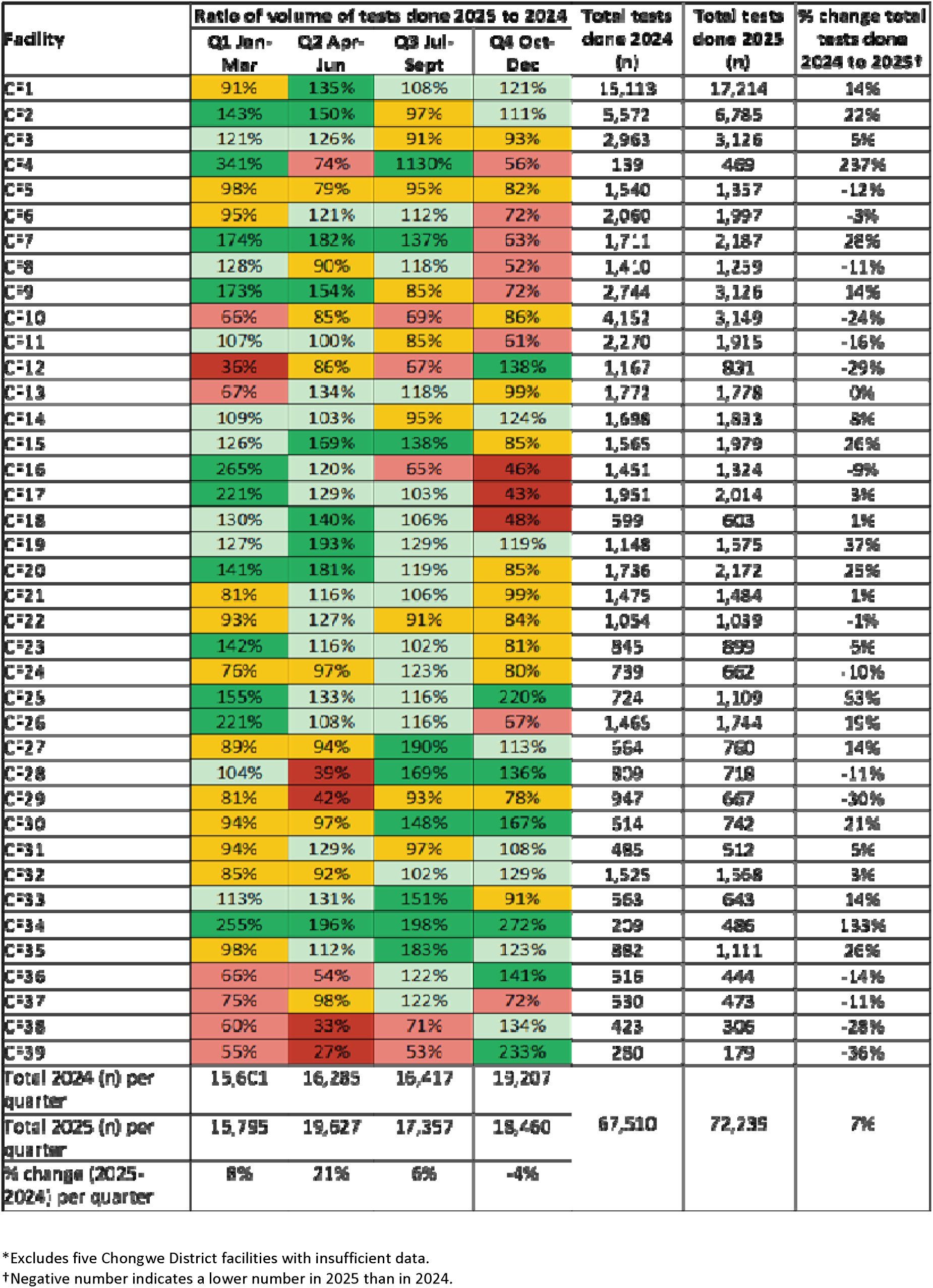
Quarterly ratio of HIV tests in 2025 relative to the corresponding quarter in 2024 in Chongwe District (CDC IP), by facility*.

Mpongwe District’s experience of HIV test volumes was very different from Chongwe’s, as shown in Figure 6. HIV testing decreased in 67% of facilities in 2025 compared to 2024. Quarterly volumes varied widely, with many sites exceeding 2024 volumes in either quarter 2 or quarter 3 and falling well below 2024 volumes in the alternate quarter. The district ended 2025 having completed 40% fewer HIV tests overall than in 2024, in contrast to Chongwe District’s net annual increase of 7% in testing volume.

**Figure 6.**
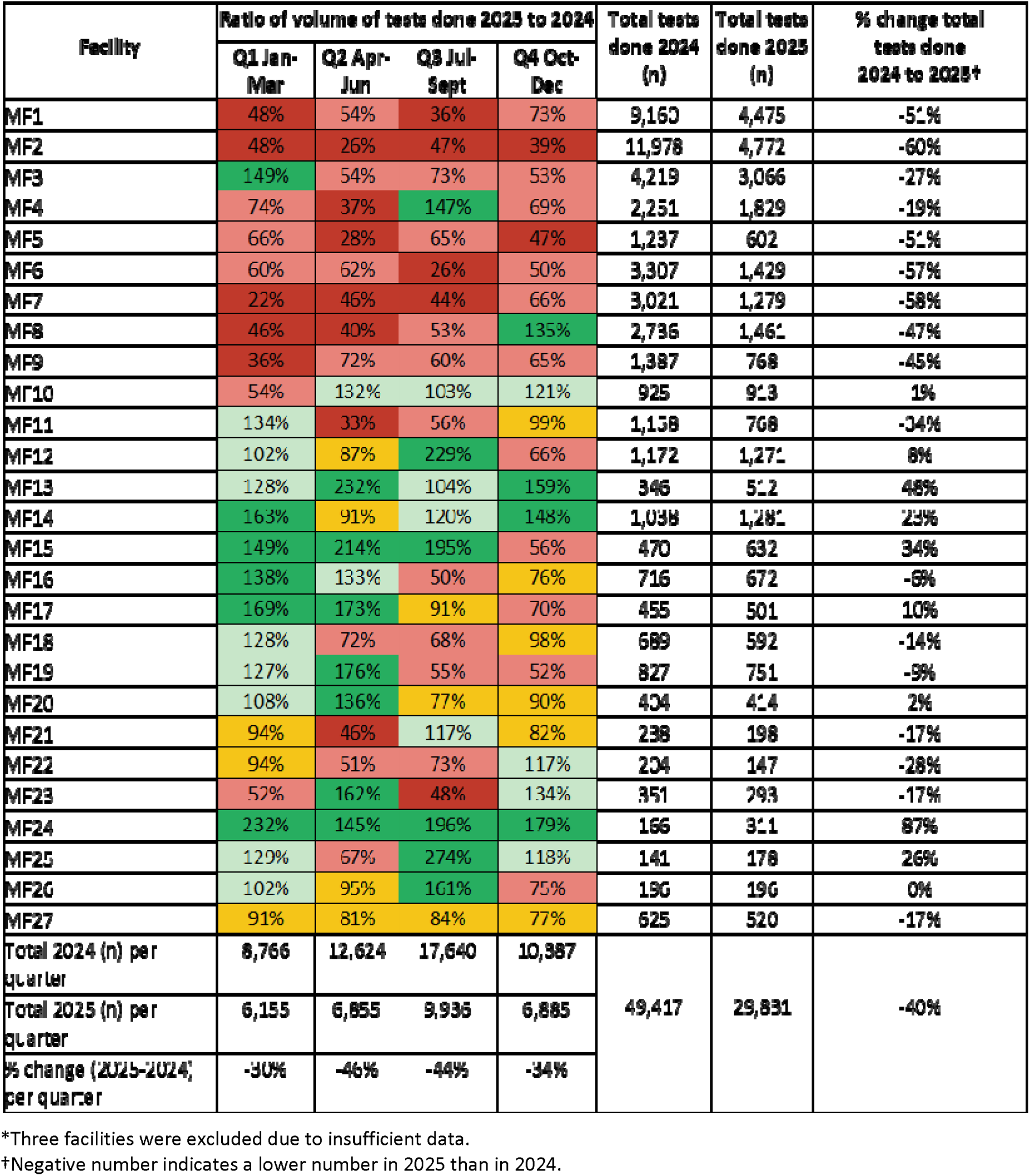
Quarterly ratio of HIV tests in 2025 relative to the corresponding quarter in 2024 in Mpongwe District (USAID IP), by facility.

### Numbers on ART in Chongwe and Mpongwe districts

After removing facilities that were missing too many quarterly observations in the database to allow analysis, the analytic dataset for numbers of clients on ART included 36 facilities in Chongwe and 25 in Mpongwe. As with the SENTINEL sites, facilities in Chongwe District reported largely stable numbers of clients on ART throughout the study period. Only 4 of the 36 sites in the district had a net loss of clients over the course of 2025, and most sites experienced gains, as shown in Figure 7.

**Figure 7.**
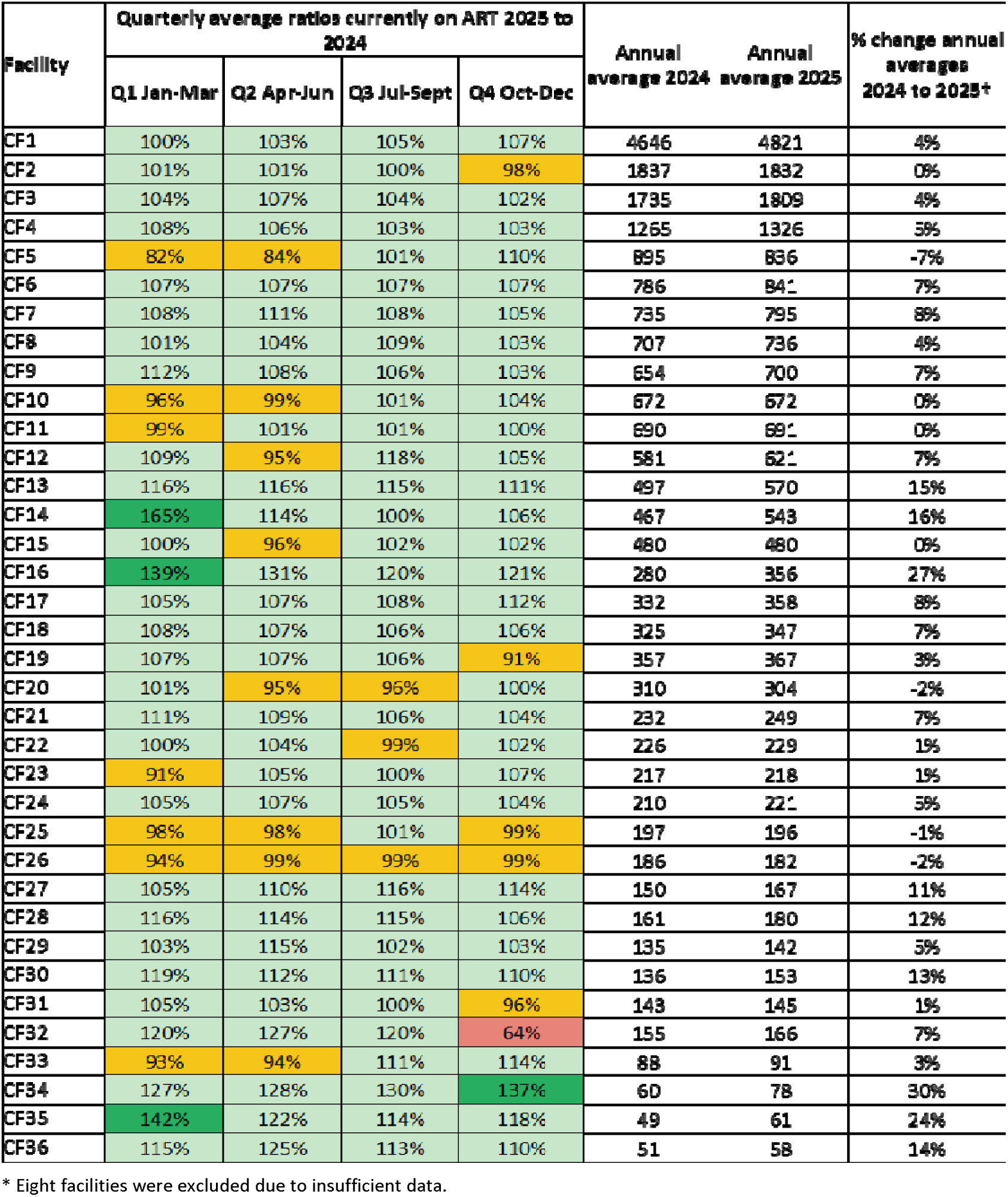
Ratios of quarterly and annual average numbers of clients on ART in 2025 relative to the corresponding interval in 2024 in Chongwe District, by facility*.

In contrast to Chongwe, facilities in Mpongwe District experienced variation in the number of clients on ART, as shown in Figure 8, with many declining by >30% in some quarters and 14 of the 25 sites suffering a net loss from 2024 to 2025. Although client numbers did grow substantially in some sites in Mpongwe District, the largest percentage increases were for the smallest facilities and represent only a handful of individual clients.

**Figure 8.**
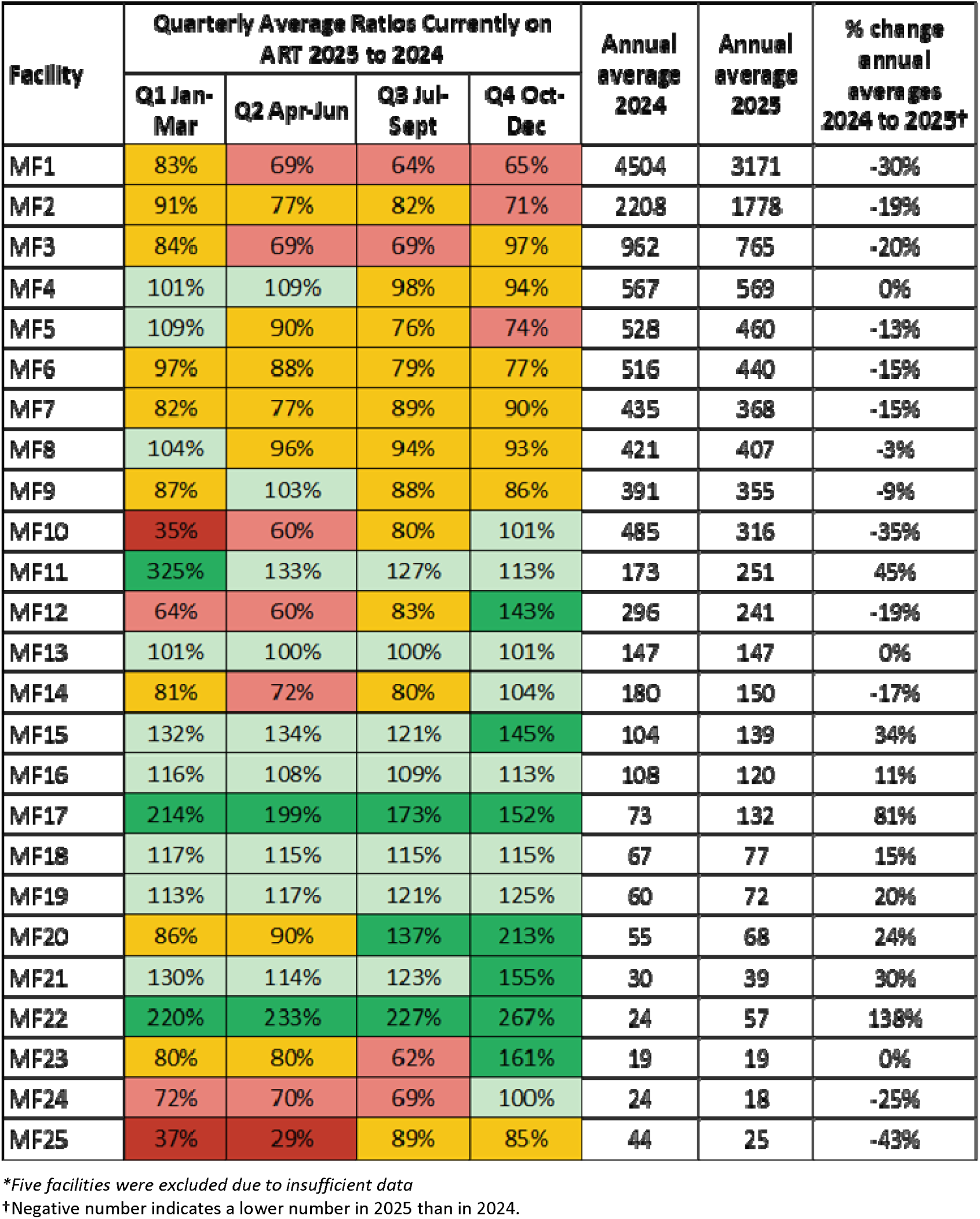
Ratios of quarterly and annual average numbers of clients on ART in 2025 relative to the corresponding interval in 2024 in Mpongwe District, by facility.

## Discussion

While negative consequences for Zambia’s HIV program were widely predicted after the 2025 PEPFAR funding disruptions, it was not anticipated how uneven both the funding cuts and the subsequent funding and service recovery would be across individual facilities. In our observational study of a sample of public health facilities in Zambia, we observed large site-level variations in these impacts. Some facilities suffered sharp reductions in the volumes of services provided, while others reported small or no changes or even increased service volumes in the months following the cuts. The results reported here suggest that both local-level and quarterly impacts may differ sharply from aggregate annual effects. The results of this study, while generally consistent with the funding experience of USAID and CDC implementing partners at the province and district level, make clear that aggregate estimates for larger geographic subdivisions and longer periods of time have missed many of the dynamics experienced by facilities in 2025.

Our primary finding was that individual facilities within the same province and/or district experienced different trajectories over the course of 2025. HIV testing volumes for 2025 both rose and fell in different quarters, compared to 2024, for most facilities in the study, though with a clear pattern of losses for formerly USAID-supported sites. ART initiations, reported only for Dataset 1, were also variable by site and province. Magnitudes of changes in ART initiations were generally smaller than for testing, but fewer recovered by the end of 2025; nearly all facilities initiated fewer clients on ART in 2025 than in 2024. Compared to testing and ART initiation volumes, in contrast, numbers of clients on ART fluctuated relatively little between years. In each province or district, some facilities ended 2025 with substantially fewer clients on ART than in 2024, while volumes of clients treated at other facilities edged up slightly between the years.

The same trend was identified in the district-level dataset (Dataset 2), where we analyzed all facilities with data available. The aggregate district-level differences between 2024 and 2025 were generally smaller in magnitude than were many of the individual facility differences. Mpongwe District suffered a 40% reduction in HIV test volumes overall, but only 7 of 27 sites in the district showed losses of more than 39%, while 8 of 27 conducted more tests in 2025 than in 2024. Similarly, Chongwe’s overall HIV testing volumes increased by 7% despite 4 of the 39 analyzed sites suffering losses of more than 25% and other sites, such as CF4 and CF34, more than doubling their testing volumes.

In considering this pattern of small net change at the district level but large gains and losses among individual facilities, we hypothesize that part of the explanation is a reallocation of clients to better-resourced facilities that maintained their service delivery capacity. The DHIS2 data used here cannot directly confirm movement of patients between facilities, and other factors such as differential restoration of staffing or commodities or facility-specific differences in completeness or accuracy of reporting may also contribute to this pattern. To the extent that reallocation does explain the observed patterns, however, clients may be making pragmatic decisions and seeking care where and when it is available. This is an important finding, as it suggests that facilities within the same small geographic area may require different types and amounts of support to recover fully from the funding cuts and that facilities that have increased service delivery volumes may also be under pressure. Presenting impacts in aggregate masks the individual impacts on health facilities, and thus limits efficacy of planning and resource allocation. The aggregate percent reduction in HIV tests across all SENTINEL sites, for example was –18%; however, within that, some sites increased their testing volumes by 25%, while in others volumes fell by as much as 45-60%. Even a district-level blanket strategy for all sites within the district may result in misallocation of resources if facility-level differences are not considered.

As implied above, the overall pattern of losses and gains in both datasets loosely reflected which U.S. government funded mechanism was in place in the province. Facilities in Central and Copperbelt provinces and Mpongwe District, where USAID supported the implementing partners and funded direct employment of clinicians, counselors, and community health workers, whose tasks included community outreach, testing in facilities and communities, appointment systems, direct clinical care for assessing and starting patients on ART, laboratory and sample courier services, defaulter tracing, and management of the EMR, experienced the most severe and most sustained reductions in service volumes. It is these direct services that Zambia’s own 2019 PEPFAR Sustainability Index and Dashboard (SID) assessment, conducted well before the 2025 funding cuts, identified as most vulnerable to disruption, in contrast to technical assistance activities such as planning, coordination, policies, governance, and quality management, which were considered less vulnerable[15]. Unlike USAID IPs, most CDC IPs (Lusaka and Southern Provinces, Chongwe District) had previously emphasized technical assistance, rather than direct service delivery, and retained some of their funding in 2025, diluting the impact of the cuts on the facilities they supported. We also note, however, multiple exceptions to this overall pattern: volumes for some services rose at some facilities previously assisted by a USAID implementing partner, while volumes for some services fell at some facilities previously (or currently) supported by a CDC IP.

Our findings are generally consistent with previous reports on the impact of the funding cuts in Zambia but are far more nuanced due to our focus on facility-level changes. Saito et al (2025)[6], looking at 48 facilities in two of Zambia’s ten provinces, saw the same sharp declines in HIV testing and ART initiation volumes we did in the first quarter of 2025 but did not report beyond March 2025 or disaggregate by facility. At the national level, PEPFAR data posted by Kenny (2026) showed an aggregate decrease of 13% in the number of individuals who were tested for HIV and received their results (HTS_TST indicator) and a 23% decrease in the number of individuals newly initiated on ART (TX_NEW indicator) between the first half of 2024 and the first half of 2025. Mulenga et al (2025)[1], modeling the likely impacts of reductions in HIV treatment access on mortality and HIV prevalence, projected large negative health impacts but also reported only at a national scale. We found no other quantitative estimates of the effects of the funding cuts, and none at facility-level.

As mentioned above, DHIS2 data alone do not reveal the reasons behind individual facilities’ losses and recoveries, beyond placing them within different IPs’ catchment areas. Several sources do, however, describe the operational impacts of the loss of IP support[1,8,16,17], providing context to our estimates of the quantitative impacts. A partial reinstatement of some of the former USAID programs through the US Department of State in mid-2025 restored some services at affected facilities[18]. Guidance from the Ministry of Health to facilities to institute task-shifting of existing government staff to cover tasks previously performed by IP staff also appears to have helped some facilities compensate for the gap [19]. Providers at individual facilities also devised their own solutions to the loss of personnel[17], which may help explain why recovery varied even among facilities supported by the same funding mechanism. Client behavior may also help explain the observed fluctuations: some potential clients may have waited until later in 2025 to seek HIV testing and/or treatment initiation services due to the loss of community outreach and the uncertainty and fear that accompanied the funding cuts[16].

This analysis had a number of potentially important limitations that should lead to cautious interpretation of findings. First and most important, the DHIS2 dataset we were able to access was incomplete and inconsistent at some facilities and for some periods. Many facilities did not have data entered for entire months or quarters, and we cannot know if these missing values represented missing data or were in fact reports of zero services delivered. Some facilities reported extremely high service delivery volumes, which could be accurate—they are not impossible--but may represent data capturing errors. We could not assess ART initiation volumes for the two district-level facility sets at all, due to missing monthly DHIS2 entries.

Second, as we speculate above, many clients may have responded to the funding cuts by seeking services at other, better-equipped facilities, and these facilities may well have been in other districts (for clients in Chongwe and Mpongwe) or in neighboring communities (for the SENTINEL sites). Our analysis did not capture these clients, making it important to differentiate between numbers of services provided (our results) and overall numbers of individuals receiving services, which our data likely underestimate. Counting of multiple events for the same client (e.g. two HIV tests for one person) is also possible, which underscores the fact that the volumes reported are for service events, not individuals.

Third, because all of our data are observational and our comparisons strictly descriptive, we can report associations only. Because our data are limited to service-delivery volumes, we could not directly measure facility-level changes in the workforce, commodity, or financing domains that the national sustainability assessment identifies as Zambia’s principal vulnerabilities; the mechanisms proposed here linking those domains to observed service disruptions remain inferential. Some of the changes observed may reflect secular developments within the Zambia HIV program or society at large. In particular, during our study period, many facilities transitioned from an older version of the SmartCare EMR (SmartCare Plus) to the current version (SmartCare Pro), resulting in data losses and data entry delays for some facilities.

Finally, our period of analysis ended in December 2025. There have been further developments among funding agencies and implementing partners as well as policy and program changes made by the Zambian Government that may limit the applicability of these findings to 2026.

## Conclusion

The 2025 disruptions to PEPFAR-supported services in Zambia were associated with highly heterogeneous changes in service delivery volumes among facilities and districts. HIV testing and ART initiation volumes fell at many facilities, especially in formerly USAID-supported areas, but increased in others. Numbers of clients on ART were generally stable, but some facilities absorbed additional clients and others saw reductions. Even within the same provinces or districts, some facilities delivered much smaller service volumes for months at a time, while neighboring facilities experienced little or no disruption.

Because of this facility-level variation in outcomes, aggregated estimates of the impacts of the funding cuts miss exactly where service delivery has been hardest hit, preventing policy makers and program managers from targeting their resources and interventions accurately. As government agencies and other stakeholders continue to take steps to compensate for the losses of donor-funded services, utilizing site-level analysis has the potential to substantially increase the efficiency of their investments and offset the uneven impacts of the funding cuts. Existing facility-level data systems such as DHIS2 make such analysis feasible in Zambia and many other countries, and the effort required to produce monthly, site-level results could be well-suited to current AI models. Providing these data in real time to those making day-to-day resource allocation decisions may be a valuable way forward.

## Supplementary files

Supplementary Table 1. Characteristics of facilities in district datasets

Supplementary Table 2. Months in which DHIS2 data were not available

## Author contributions

Conceptualization: MM, SR, AH, IH

Data Curation: MM, TN, CM

Formal Analysis: MM, SR, MB

Funding Acquisition: SR

Methodology: MM, SR

Project Administration: TN, PH, HS

Resources: SS

Supervision: SR, TN, PH, AH, IM

Visualization: MM, SR, AM

Writing – Original Draft Preparation: MM, SR, IH

Writing – Review & Editing: All co-authors

## Funding

Funding for the study was provided by the Gates Foundation through INV-088605 to the Wits Health Consortium. The funder had no role in study design, data collection and analysis, decision to publish, or preparation of the manuscript.

## Competing interests

The authors declare no competing interests. SS and CM are employed by the government agency that has supervisory authority over the study sites.

## Data availability

All data used for this study are owned by the Zambia Ministry The facility-level data analyzed in this study were obtained from Zambia’s Ministry of Health DHIS2 system and are not publicly available from the authors. Researchers seeking access should request permission from the Zambia Ministry of Health at, specifying the indicators, facilities, time period, and intended use. Access is subject to Ministry approval.

---

**Supplementary Table 1.** Characteristics of facilities in district datasets.

| Facility | District | Setting |
| --- | --- | --- |
| CF1 | Chongwe | Rural |
| CF2 | Chongwe | Rural |
| CF3 | Chongwe | Urban |
| CF4 | Chongwe | Rural |
| CF5 | Chongwe | Rural |
| CF6 | Chongwe | Rural |
| CF7 | Chongwe | Rural |
| CF8 | Chongwe | Rural |
| CF9 | Chongwe | Urban |
| CF10 | Chongwe | Rural |
| CF11 | Chongwe | Rural |
| CF12 | Chongwe | Rural |
| CF13 | Chongwe | Rural |
| CF14 | Chongwe | Rural |
| CF15 | Chongwe | Rural |
| CF16 | Chongwe | Rural |
| CF17 | Chongwe | Rural |
| CF18 | Chongwe | Rural |
| CF19 | Chongwe | Rural |
| CF20 | Chongwe | Rural |
| CF21 | Chongwe | Rural |
| CF22 | Chongwe | Rural |
| CF23 | Chongwe | Rural |
| CF24 | Chongwe | Rural |
| CF25 | Chongwe | Rural |
| CF26 | Chongwe | Rural |
| CF27 | Chongwe | Rural |
| CF28 | Chongwe | Rural |
| CF29 | Chongwe | Rural |
| CF30 | Chongwe | Rural |
| CF31 | Chongwe | Rural |
| CF32 | Chongwe | Rural |
| CF33 | Chongwe | Rural |
| CF34 | Chongwe | Rural |
| CF35 | Chongwe | Rural |
| CF36 | Chongwe | Urban |
| CF37 | Chongwe | Rural |
| CF38 | Chongwe | Rural |
| CF39 | Chongwe | Rural |
| CF40 | Chongwe | Rural |
| CF41 | Chongwe | Rural |
| CF42 | Chongwe | Rural |
| CF43 | Chongwe | Urban |
| MF1 | Mpongwe | Rural |
| MF2 | Mpongwe | Rural |
| MF3 | Mpongwe | Rural |
| MF4 | Mpongwe | Rural |
| MF5 | Mpongwe | Rural |
| MF6 | Mpongwe | Rural |
| MF7 | Mpongwe | Rural |
| MF8 | Mpongwe | Rural |
| MF9 | Mpongwe | Rural |
| MF10 | Mpongwe | Rural |
| MF11 | Mpongwe | Rural |
| MF12 | Mpongwe | Rural |
| MF13 | Mpongwe | Rural |
| MF14 | Mpongwe | Rural |
| MF15 | Mpongwe | Rural |
| MF16 | Mpongwe | Rural |
| MF17 | Mpongwe | Rural |
| MF18 | Mpongwe | Rural |
| MF19 | Mpongwe | Rural |
| MF20 | Mpongwe | Rural |
| MF21 | Mpongwe | Rural |
| MF22 | Mpongwe | Rural |
| MF23 | Mpongwe | Rural |
| MF24 | Mpongwe | Rural |
| MF25 | Mpongwe | Rural |
| MF26 | Mpongwe | Rural |
| MF27 | Mpongwe | Rural |
| MF28 | Mpongwe | Rural |
| MF29 | Mpongwe | Rural |
| MF30 | Mpongwe | Rural |

**Supplementary Table 2a.**
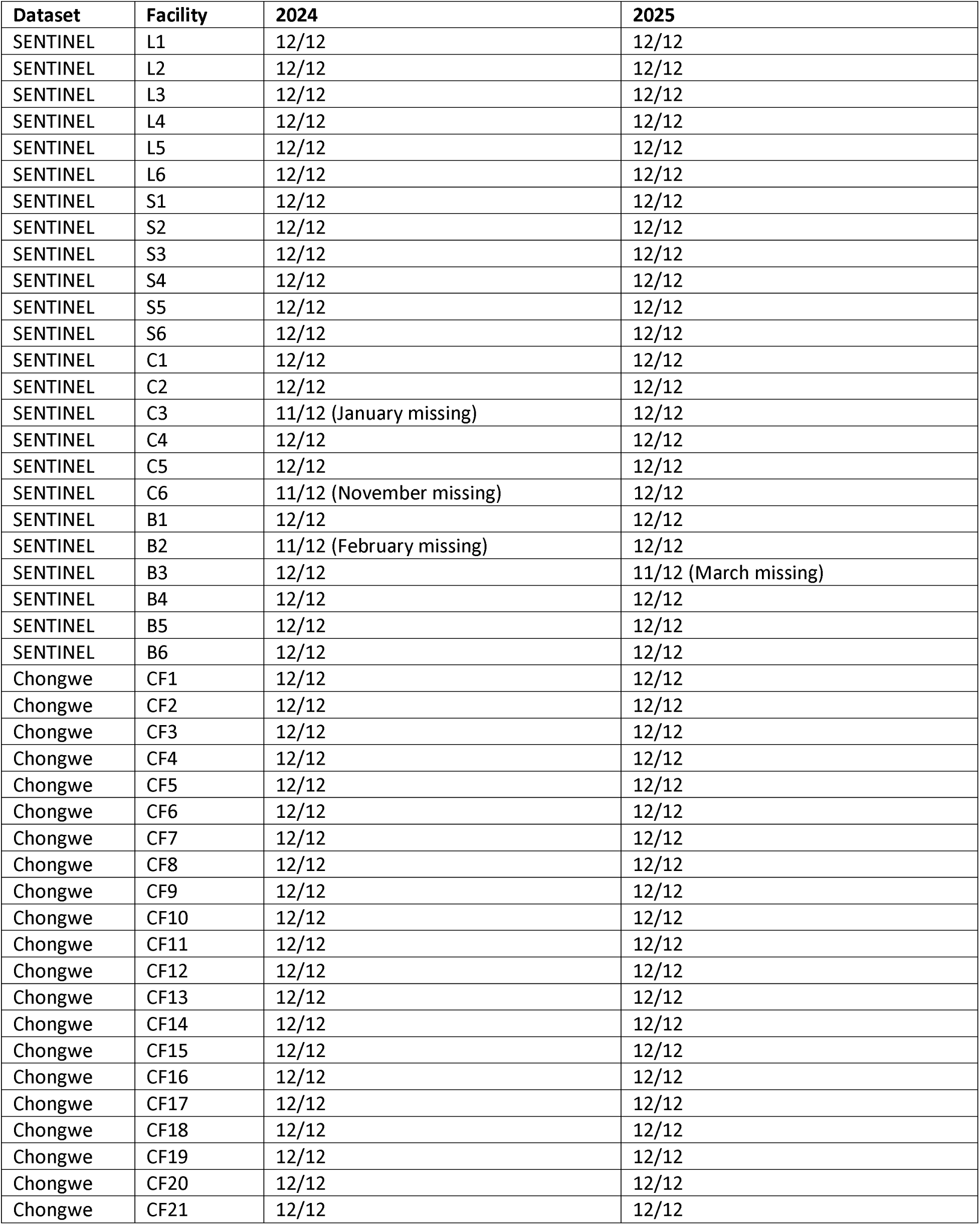
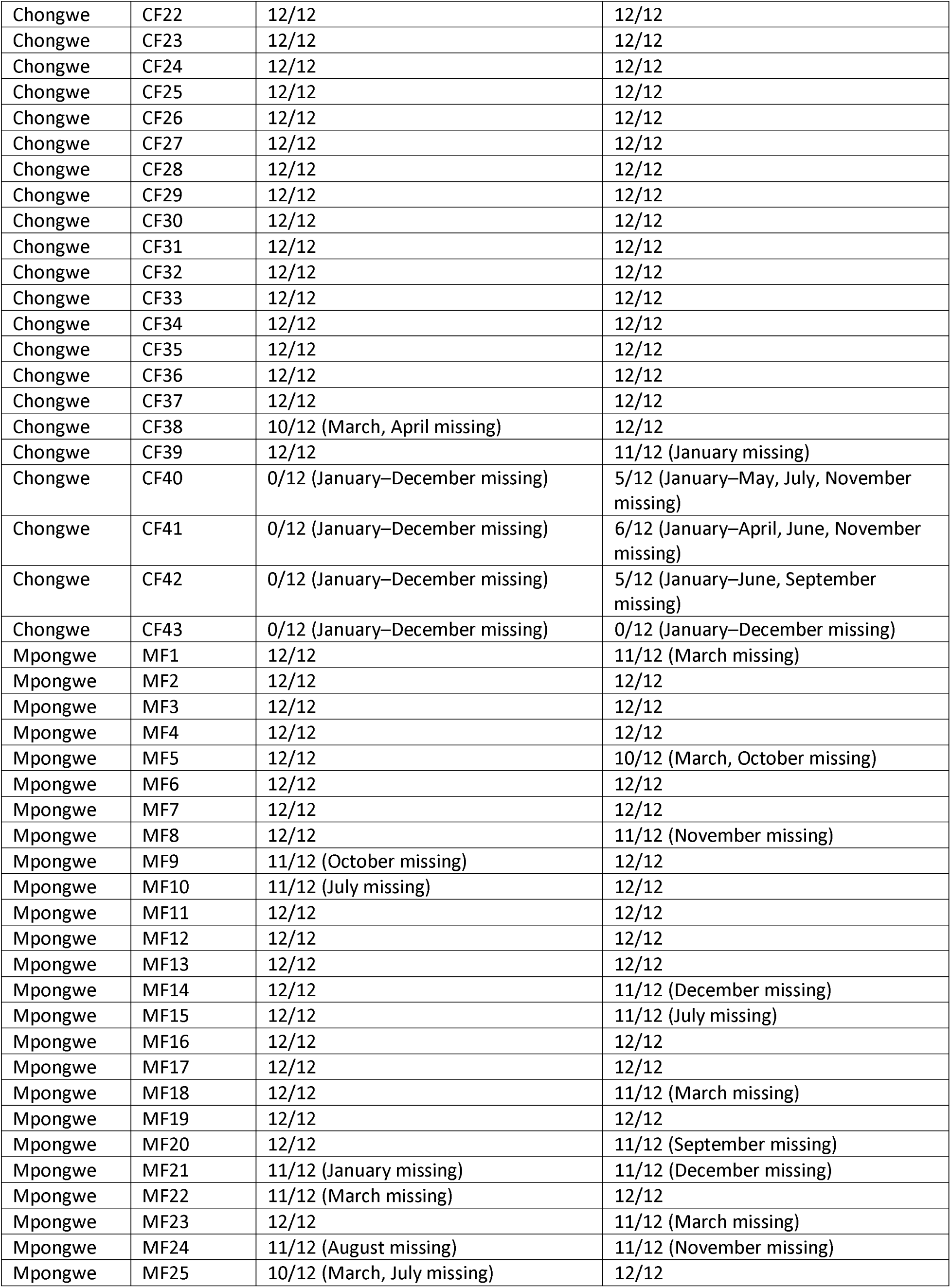
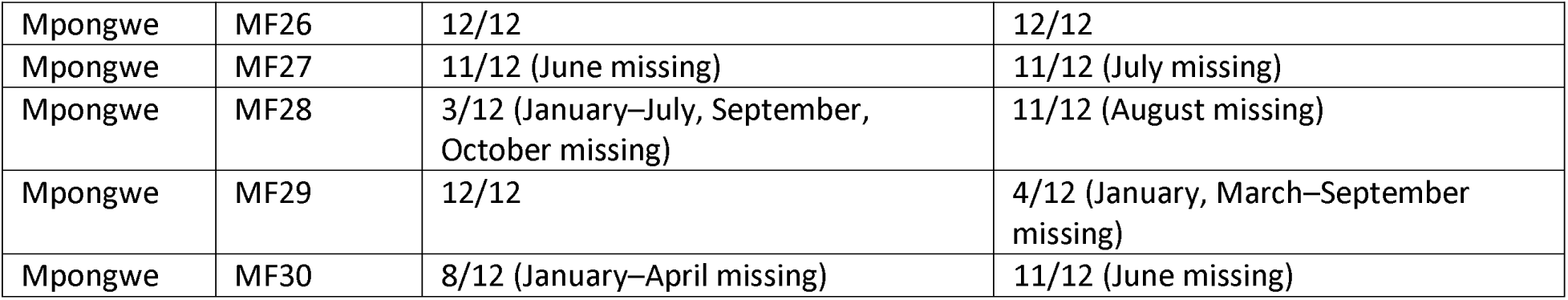
HIV testing volumes: months in which data were missing and the number of months contributing data, by facility and quarter.

| Dataset | Facility | 2024 | 2025 |
| --- | --- | --- | --- |
| SENTINEL | L1 | 12/12 | 12/12 |
| SENTINEL | L2 | 12/12 | 12/12 |
| SENTINEL | L3 | 12/12 | 12/12 |
| SENTINEL | L4 | 12/12 | 12/12 |
| SENTINEL | L5 | 12/12 | 12/12 |
| SENTINEL | L6 | 12/12 | 12/12 |
| SENTINEL | S1 | 12/12 | 12/12 |
| SENTINEL | S2 | 12/12 | 12/12 |
| SENTINEL | S3 | 12/12 | 12/12 |
| SENTINEL | S4 | 12/12 | 12/12 |
| SENTINEL | S5 | 12/12 | 12/12 |
| SENTINEL | S6 | 12/12 | 12/12 |
| SENTINEL | C1 | 12/12 | 12/12 |
| SENTINEL | C2 | 12/12 | 12/12 |
| SENTINEL | C3 | 11/12 (January missing) | 12/12 |
| SENTINEL | C4 | 12/12 | 12/12 |
| SENTINEL | C5 | 12/12 | 12/12 |
| SENTINEL | C6 | 11/12 (November missing) | 12/12 |
| SENTINEL | B1 | 12/12 | 12/12 |
| SENTINEL | B2 | 11/12 (February missing) | 12/12 |
| SENTINEL | B3 | 12/12 | 11/12 (March missing) |
| SENTINEL | B4 | 12/12 | 12/12 |
| SENTINEL | B5 | 12/12 | 12/12 |
| SENTINEL | B6 | 12/12 | 12/12 |
| Chongwe | CF1 | 12/12 | 12/12 |
| Chongwe | CF2 | 12/12 | 12/12 |
| Chongwe | CF3 | 12/12 | 12/12 |
| Chongwe | CF4 | 12/12 | 12/12 |
| Chongwe | CF5 | 12/12 | 12/12 |
| Chongwe | CF6 | 12/12 | 12/12 |
| Chongwe | CF7 | 12/12 | 12/12 |
| Chongwe | CF8 | 12/12 | 12/12 |
| Chongwe | CF9 | 12/12 | 12/12 |
| Chongwe | CF10 | 12/12 | 12/12 |
| Chongwe | CF11 | 12/12 | 12/12 |
| Chongwe | CF12 | 12/12 | 12/12 |
| Chongwe | CF13 | 12/12 | 12/12 |
| Chongwe | CF14 | 12/12 | 12/12 |
| Chongwe | CF15 | 12/12 | 12/12 |
| Chongwe | CF16 | 12/12 | 12/12 |
| Chongwe | CF17 | 12/12 | 12/12 |
| Chongwe | CF18 | 12/12 | 12/12 |
| Chongwe | CF19 | 12/12 | 12/12 |
| Chongwe | CF20 | 12/12 | 12/12 |
| Chongwe | CF21 | 12/12 | 12/12 |

| **Dataset** | **Facility** | **2024** | **2025** |
| --- | --- | --- | --- |
| Chongwe | CF22 | 12/12 | 12/12 |
| Chongwe | CF23 | 12/12 | 12/12 |
| Chongwe | CF24 | 12/12 | 12/12 |
| Chongwe | CF25 | 12/12 | 12/12 |
| Chongwe | CF26 | 12/12 | 12/12 |
| Chongwe | CF27 | 12/12 | 12/12 |
| Chongwe | CF28 | 12/12 | 12/12 |
| Chongwe | CF29 | 12/12 | 12/12 |
| Chongwe | CF30 | 12/12 | 12/12 |
| Chongwe | CF31 | 12/12 | 12/12 |
| Chongwe | CF32 | 12/12 | 12/12 |
| Chongwe | CF33 | 12/12 | 12/12 |
| Chongwe | CF34 | 12/12 | 12/12 |
| Chongwe | CF35 | 12/12 | 12/12 |
| Chongwe | CF36 | 12/12 | 12/12 |
| Chongwe | CF37 | 12/12 | 12/12 |
| Chongwe | CF38 | 10/12 (March, April missing) | 12/12 |
| Chongwe | CF39 | 12/12 | 11/12 (January missing) |
| Chongwe | CF40 | 0/12 (January–December missing) | 5/12 (January–May, July, November missing) |
| Chongwe | CF41 | 0/12 (January–December missing) | 6/12 (January–April, June, November missing) |
| Chongwe | CF42 | 0/12 (January–December missing) | 5/12 (January–June, September missing) |
| Chongwe | CF43 | 0/12 (January–December missing) | 0/12 (January–December missing) |
| Mpongwe | MF1 | 12/12 | 11/12 (March missing) |
| Mpongwe | MF2 | 12/12 | 12/12 |
| Mpongwe | MF3 | 12/12 | 12/12 |
| Mpongwe | MF4 | 12/12 | 12/12 |
| Mpongwe | MF5 | 12/12 | 10/12 (March, October missing) |
| Mpongwe | MF6 | 12/12 | 12/12 |
| Mpongwe | MF7 | 12/12 | 12/12 |
| Mpongwe | MF8 | 12/12 | 11/12 (November missing) |
| Mpongwe | MF9 | 11/12 (October missing) | 12/12 |
| Mpongwe | MF10 | 11/12 (July missing) | 12/12 |
| Mpongwe | MF11 | 12/12 | 12/12 |
| Mpongwe | MF12 | 12/12 | 12/12 |
| Mpongwe | MF13 | 12/12 | 12/12 |
| Mpongwe | MF14 | 12/12 | 11/12 (December missing) |
| Mpongwe | MF15 | 12/12 | 11/12 (July missing) |
| Mpongwe | MF16 | 12/12 | 12/12 |
| Mpongwe | MF17 | 12/12 | 12/12 |
| Mpongwe | MF18 | 12/12 | 11/12 (March missing) |
| Mpongwe | MF19 | 12/12 | 12/12 |
| Mpongwe | MF20 | 12/12 | 11/12 (September missing) |
| Mpongwe | MF21 | 11/12 (January missing) | 11/12 (December missing) |
| Mpongwe | MF22 | 11/12 (March missing) | 12/12 |
| Mpongwe | MF23 | 12/12 | 11/12 (March missing) |
| Mpongwe | MF24 | 11/12 (August missing) | 11/12 (November missing) |
| Mpongwe | MF25 | 10/12 (March, July missing) | 12/12 |
| Mpongwe | MF26 | 12/12 | 12/12 |
| Mpongwe | MF27 | 11/12 (June missing) | 11/12 (July missing) |
| Mpongwe | MF28 | 3/12 (January–July, September, October missing) | 11/12 (August missing) |
| Mpongwe | MF29 | 12/12 | 4/12 (January, March–September missing) |
| Mpongwe | MF30 | 8/12 (January–April missing) | 11/12 (June missing) |

**Supplementary Table 2b.**
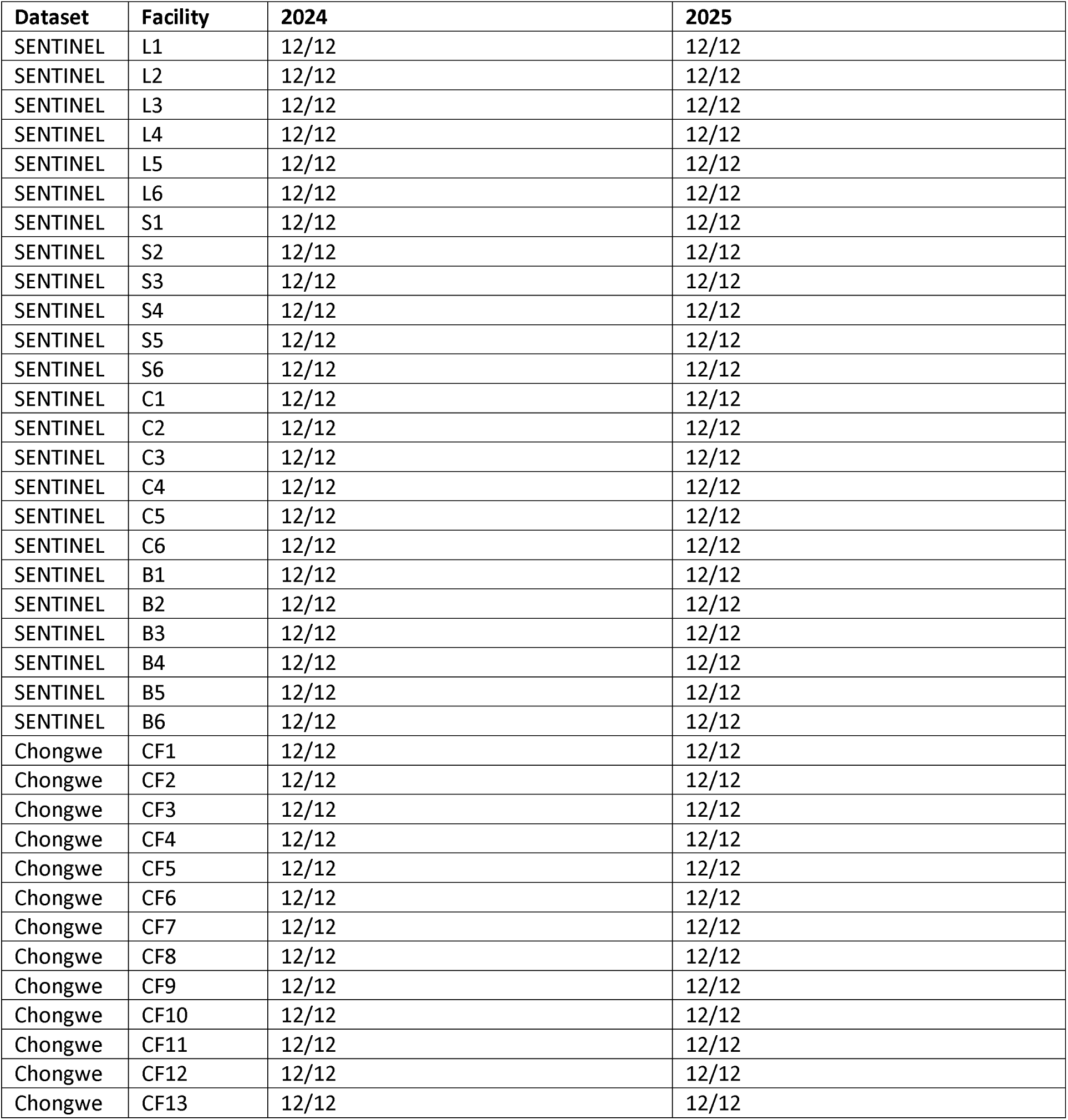
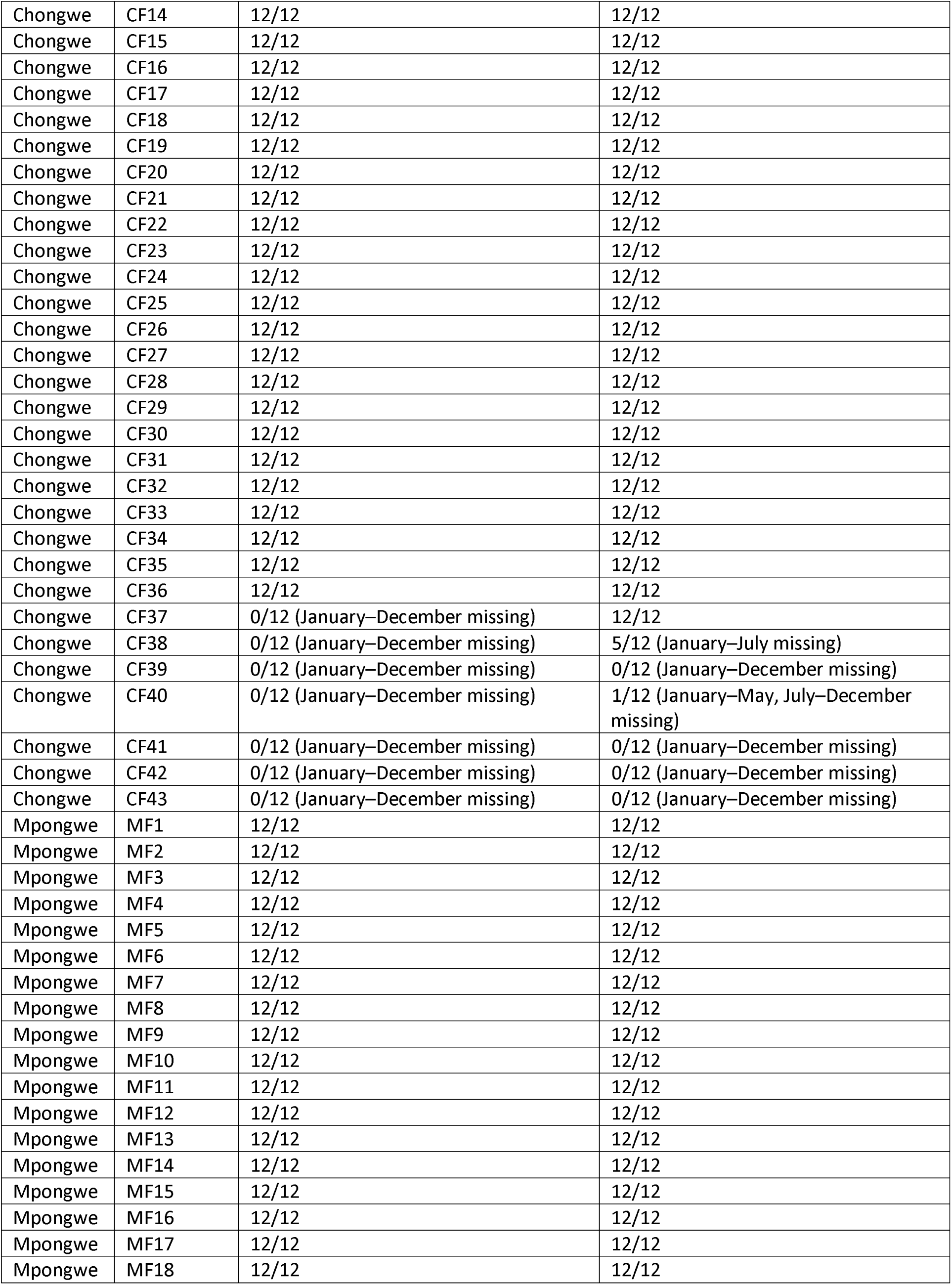
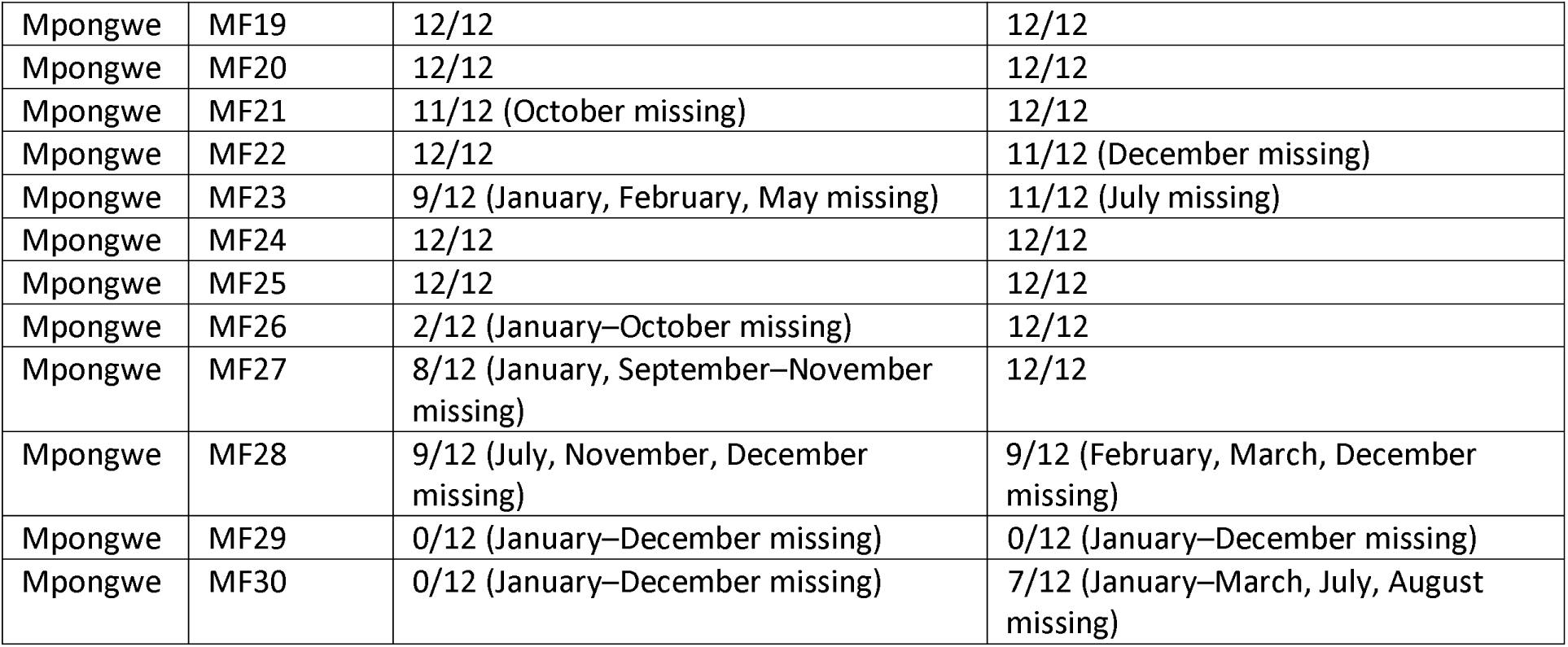
Current numbers on ART: months in which data were missing and the number of months contributing data, by facility and year.

| Dataset | Facility | 2024 | 2025 |
| --- | --- | --- | --- |
| SENTINEL | L1 | 12/12 | 12/12 |
| SENTINEL | L2 | 12/12 | 12/12 |
| SENTINEL | L3 | 12/12 | 12/12 |
| SENTINEL | L4 | 12/12 | 12/12 |
| SENTINEL | L5 | 12/12 | 12/12 |
| SENTINEL | L6 | 12/12 | 12/12 |
| SENTINEL | S1 | 12/12 | 12/12 |
| SENTINEL | S2 | 12/12 | 12/12 |
| SENTINEL | S3 | 12/12 | 12/12 |
| SENTINEL | S4 | 12/12 | 12/12 |
| SENTINEL | S5 | 12/12 | 12/12 |
| SENTINEL | S6 | 12/12 | 12/12 |
| SENTINEL | C1 | 12/12 | 12/12 |
| SENTINEL | C2 | 12/12 | 12/12 |
| SENTINEL | C3 | 12/12 | 12/12 |
| SENTINEL | C4 | 12/12 | 12/12 |
| SENTINEL | C5 | 12/12 | 12/12 |
| SENTINEL | C6 | 12/12 | 12/12 |
| SENTINEL | B1 | 12/12 | 12/12 |
| SENTINEL | B2 | 12/12 | 12/12 |
| SENTINEL | B3 | 12/12 | 12/12 |
| SENTINEL | B4 | 12/12 | 12/12 |
| SENTINEL | B5 | 12/12 | 12/12 |
| SENTINEL | B6 | 12/12 | 12/12 |
| Chongwe | CF1 | 12/12 | 12/12 |
| Chongwe | CF2 | 12/12 | 12/12 |
| Chongwe | CF3 | 12/12 | 12/12 |
| Chongwe | CF4 | 12/12 | 12/12 |
| Chongwe | CF5 | 12/12 | 12/12 |
| Chongwe | CF6 | 12/12 | 12/12 |
| Chongwe | CF7 | 12/12 | 12/12 |
| Chongwe | CF8 | 12/12 | 12/12 |
| Chongwe | CF9 | 12/12 | 12/12 |
| Chongwe | CF10 | 12/12 | 12/12 |
| Chongwe | CF11 | 12/12 | 12/12 |
| Chongwe | CF12 | 12/12 | 12/12 |
| Chongwe | CF13 | 12/12 | 12/12 |

| **Dataset** | **Facility** | **2024** | **2025** |
| --- | --- | --- | --- |
| Chongwe | CF14 | 12/12 | 12/12 |
| Chongwe | CF15 | 12/12 | 12/12 |
| Chongwe | CF16 | 12/12 | 12/12 |
| Chongwe | CF17 | 12/12 | 12/12 |
| Chongwe | CF18 | 12/12 | 12/12 |
| Chongwe | CF19 | 12/12 | 12/12 |
| Chongwe | CF20 | 12/12 | 12/12 |
| Chongwe | CF21 | 12/12 | 12/12 |
| Chongwe | CF22 | 12/12 | 12/12 |
| Chongwe | CF23 | 12/12 | 12/12 |
| Chongwe | CF24 | 12/12 | 12/12 |
| Chongwe | CF25 | 12/12 | 12/12 |
| Chongwe | CF26 | 12/12 | 12/12 |
| Chongwe | CF27 | 12/12 | 12/12 |
| Chongwe | CF28 | 12/12 | 12/12 |
| Chongwe | CF29 | 12/12 | 12/12 |
| Chongwe | CF30 | 12/12 | 12/12 |
| Chongwe | CF31 | 12/12 | 12/12 |
| Chongwe | CF32 | 12/12 | 12/12 |
| Chongwe | CF33 | 12/12 | 12/12 |
| Chongwe | CF34 | 12/12 | 12/12 |
| Chongwe | CF35 | 12/12 | 12/12 |
| Chongwe | CF36 | 12/12 | 12/12 |
| Chongwe | CF37 | 0/12 (January–December missing) | 12/12 |
| Chongwe | CF38 | 0/12 (January–December missing) | 5/12 (January–July missing) |
| Chongwe | CF39 | 0/12 (January–December missing) | 0/12 (January–December missing) |
| Chongwe | CF40 | 0/12 (January–December missing) | 1/12 (January–May, July–December missing) |
| Chongwe | CF41 | 0/12 (January–December missing) | 0/12 (January–December missing) |
| Chongwe | CF42 | 0/12 (January–December missing) | 0/12 (January–December missing) |
| Chongwe | CF43 | 0/12 (January–December missing) | 0/12 (January–December missing) |
| Mpongwe | MF1 | 12/12 | 12/12 |
| Mpongwe | MF2 | 12/12 | 12/12 |
| Mpongwe | MF3 | 12/12 | 12/12 |
| Mpongwe | MF4 | 12/12 | 12/12 |
| Mpongwe | MF5 | 12/12 | 12/12 |
| Mpongwe | MF6 | 12/12 | 12/12 |
| Mpongwe | MF7 | 12/12 | 12/12 |
| Mpongwe | MF8 | 12/12 | 12/12 |
| Mpongwe | MF9 | 12/12 | 12/12 |
| Mpongwe | MF10 | 12/12 | 12/12 |
| Mpongwe | MF11 | 12/12 | 12/12 |
| Mpongwe | MF12 | 12/12 | 12/12 |
| Mpongwe | MF13 | 12/12 | 12/12 |
| Mpongwe | MF14 | 12/12 | 12/12 |
| Mpongwe | MF15 | 12/12 | 12/12 |
| Mpongwe | MF16 | 12/12 | 12/12 |
| Mpongwe | MF17 | 12/12 | 12/12 |
| Mpongwe | MF18 | 12/12 | 12/12 |
| Mpongwe | MF19 | 12/12 | 12/12 |
| Mpongwe | MF20 | 12/12 | 12/12 |
| Mpongwe | MF21 | 11/12 (October missing) | 12/12 |
| Mpongwe | MF22 | 12/12 | 11/12 (December missing) |
| Mpongwe | MF23 | 9/12 (January, February, May missing) | 11/12 (July missing) |
| Mpongwe | MF24 | 12/12 | 12/12 |
| Mpongwe | MF25 | 12/12 | 12/12 |
| Mpongwe | MF26 | 2/12 (January–October missing) | 12/12 |
| Mpongwe | MF27 | 8/12 (January, September–November missing) | 12/12 |
| Mpongwe | MF28 | 9/12 (July, November, December missing) | 9/12 (February, March, December missing) |
| Mpongwe | MF29 | 0/12 (January–December missing) | 0/12 (January–December missing) |
| Mpongwe | MF30 | 0/12 (January–December missing) | 7/12 (January–March, July, August missing) |

**Supplementary Table 2C.** Treatment Initiations: months in which data were missing and the number of months contributing data, by facility and year, SENTINEL site.

| Dataset | Facility | 2024 | 2025 |
| --- | --- | --- | --- |
| SENTINEL | L1 | 12/12 | 12/12 |
| SENTINEL | L2 | 12/12 | 12/12 |
| SENTINEL | L3 | 12/12 | 12/12 |
| SENTINEL | L4 | 12/12 | 10/12 |
| SENTINEL | L5 | 12/12 | 12/12 |
| SENTINEL | L6 | 12/12 | 12/12 |
| SENTINEL | S1 | 12/12 | 12/12 |
| SENTINEL | S2 | 12/12 | 12/12 |
| SENTINEL | S3 | 12/12 | 10/12 |
| SENTINEL | S4 | 12/12 | 12/12 |
| SENTINEL | S5 | 12/12 | 12/12 |
| SENTINEL | S6 | 12/12 | 12/12 |
| SENTINEL | C1 | 12/12 | 12/12 |
| SENTINEL | C2 | 12/12 | 10/12 |
| SENTINEL | C3 | 12/12 | 12/12 |
| SENTINEL | C4 | 12/12 | 12/12 |
| SENTINEL | C5 | 12/12 | 12/12 |
| SENTINEL | C6 | 12/12 | 12/12 |
| SENTINEL | B1 | 12/12 | 10/12 |
| SENTINEL | B2 | 12/12 | 12/12 |
| SENTINEL | B3 | 12/12 | 11/12 (December missing) |
| SENTINEL | B4 | 12/12 | 12/12 |
| SENTINEL | B5 | 12/12 | 12/12 |
| SENTINEL | B6 | 11/12 (September missing) | 12/12 |

## Notes

### Author Declarations

Access to DHIS2 data was approved by the ERES Converge IRB through the GREAT protocol (2019-Sep-030) and the SHIFT protocol (2025-Sep-011). Both the GREAT and SHIFT protocols were also approved by the National Health Research Authority (NHRA) in Zambia and by the Boston University Medical Campus IRB (GREAT: H-38823; SHIFT: H-46287).

